# Effectiveness of interventions involving parents on children’s eating behaviours: protocol for a systematic review and meta-analysis

**DOI:** 10.1101/2025.10.06.25337454

**Authors:** Fatemeh Khorramrouz, Sarah Rae, Michaela Kucab, Elizabeth M. Uleryk, Jonathon L Maguire, Petros Pechlivanoglou, Kevin Thorpe, Elena M Comelli, Catherine S. Birken TARGet Kids! collaboration

**Affiliations:** Department of Nutritional Sciences, University of Toronto, Toronto, Ontario, Canada; Child Health Evaluative Sciences, SickKids Research Institute, Toronto, Ontario, Canada; Edwin S.H. Leong Centre for Healthy Children, University of Toronto, Toronto, Ontario, Canada; Institute of Health Policy Management and Evaluation, University of Toronto, Toronto, Ontario, Canada; Joannah & Brian Lawson Centre for Child Nutrition, University of Toronto, Toronto, Ontario, Canada; Department of Pediatrics, St Michael’s Hospital, Toronto, Ontario, Canada; EM Uleryk Consulting, Mississauga, ON, Canada; Li Ka Shing Knowledge Institute, St. Michael’s Hospital, Toronto Ontario Canada; Dalla Lana School of Public Health, University of Toronto, Toronto, Ontario, Canada; Department of Pediatrics, Faculty of Medicine, University of Toronto, Toronto, Ontario, Canada; SickKids Research Institute, Toronto, Ontario, Canada; Division of Pediatric Medicine, Hospital for Sick Children, Toronto, Ontario, Canada

**Keywords:** Eating Behaviours, Children, Parents, Randomized Controlled Trial, Systematic Review, Meta-analysis

## Abstract

**Background:** Parents play a pivotal role in shaping their children’s food environment and eating behaviours. Involving parents in interventions designed to promote nutritional outcomes, such as dietary intake in children has been shown to improve parental feeding practices. However, it remains unclear how such interventions influence children’s eating behaviour outcomes. This protocol describes the methods of a systematic review evaluating the effectiveness of interventions involving parents in improving the eating behaviours of healthy children aged 0 to 12 years.

**Methods and analysis:** Electronic databases including MEDLINE, EMBASE, CENTRAL, APA PsycINFO, CINAHL, Scopus, and Web of Science will be searched from inception to September 2025. A search strategy is developed to identify randomized controlled trials (RCTs) directly involving parents and reporting eating behaviours in children as either primary or secondary outcomes. Two independent reviewers will screen identified records and extract data on study, participant and intervention characteristics. Study results relevant to our primary and secondary outcomes will also be extracted using a pre-piloted standardized data extraction form. We will use the Cochrane Risk of Bias tool and GRADE approach to assess risk of bias and quality of evidence assessment, respectively. Where possible, meta-analysis using random-effects models will be performed, otherwise a qualitative summary will be provided.

**Ethics and dissemination:** Ethics approval is not required for this study as no primary data will be collected. The findings will provide valuable insights for stakeholders to inform and optimize public health policies and practices aimed at empowering families to promote healthy eating behaviours early in childhood. The results will be submitted for publication in a peer-reviewed journal.

**PROSPERO registration number:** CRD420251076540

**Strengths and limitations of this study:**

- This review will include only randomized controlled trials, which are considered the gold standard for evaluating intervention effectiveness and provide high-quality evidence.
- A comprehensive search strategy across multiple electronic databases and grey literature will be used to identify the existing literature.
- Study selection, data extraction, and risk of bias assessment will be conducted independently by two reviewers to enhance reliability and reduce potential bias.
- Variability in intervention components, outcome measures, and follow-up durations may introduce heterogeneity and limit the interpretability of findings.
- Excluding non-randomized studies may result in the omission of potentially relevant evidence from real-world or community-based interventions that lack trial-based designs.

## Introduction

Childhood is an important period for the development of eating behaviours(1). These behaviours can be broadly categorized into food-approach behaviours—such as food responsiveness and emotional overeating—which are linked to higher energy intake and risk of obesity, and food-avoidant behaviours—such as satiety responsiveness and food fussiness—which are associated with lower energy intake and risk of underweight and nutritional deficiencies(2–4). Poor eating behaviours formed during childhood may impair children’s sensitivity to internal signals of satiety and hunger(5), which can negatively impact dietary intake(6), cardiometabolic health(7), and overall growth and development (8). Evidence also suggests that eating behaviours established early in life often persist into adulthood, increasing the risk of chronic diseases such as obesity(4).

As primary caregivers, parents play a pivotal role in shaping their children’s feeding environment and eating experiences(9). Throughout childhood, parents are the primary food providers and gatekeepers, regulating what, when, and how much their children eat(9). Given this influence, parenting skill development and feeding practices have become central components of interventions aimed at fostering healthy behaviours in children(10). Parent-involved interventions are structured programs that actively engage parents to improve child health and development by equipping them with skills and strategies to create a healthy home environment and support their child’s behaviour and well-being(11).

Previous reviews of intervention studies have primarily focused on the impact of parent-based interventions on children’s dietary intake and weight-related outcomes(10,12,13). A 2015 systematic review and meta-analysis of eight randomised clinical trials (RCTs) involving 1,487 parent-child dyads evaluated parent-based interventions targeting obesity prevention and treatment among children aged 5 to 15 years(14). While effects on children’s BMI or BMI z-score were not found, some improvements were observed in dietary outcomes, such as reduced intake of energy-dense foods. More recently, a 2024 systematic review of 12 studies focused on parenting interventions delivered electronically (eHealth) for children aged 3–6 years(12). The interventions showed significant improvements in dietary behaviours, including increased fruit and vegetable consumption and reduced intake of sugar-sweetened beverages. However, these reviews did not examine eating behaviour outcomes. This remains an important knowledge gap, as early childhood is a sensitive developmental period when parents play a central role in shaping children’s eating behaviours and food environments.

This systematic review will focus on children aged 0 to 12 years, as they are highly dependent on parental guidance before transitioning to adolescence. Understanding the effectiveness of interventions involving parents in shaping children’s eating behaviours is essential for designing evidence-based strategies that can be implemented across diverse settings. This review will address a critical gap in the literature and provide actionable evidence for policymakers, health practitioners, and researchers working to promote healthy eating behaviours early in life through active parental involvement.

### Objectives

The primary objective of this systematic review will be to evaluate the effects of parent-involved interventions on eating behaviours of healthy children aged 0 to 12 years. Secondary objectives are to assess the effects of these interventions on children’s dietary intake, anthropometric outcomes, quality of life, and parental feeding practices. The characteristics and specific features associated with the effectiveness of various intervention programs will also be examined.

## Methods

### Design

This systematic review protocol was designed according to the Preferred Reporting Items for Systematic Reviews and Meta-Analysis Protocols 2015 (PRISMA-P) statement (15). A completed PRISMA-P checklist is provided in **supplementary Data 1**. If any changes to the protocol are required, we will document the date of the change, details of the amendment, and rationale within PROSPERO. We will also mention the amendment(s) within the final manuscript of results.

### Eligibility criteria

The inclusion of studies for this review will be based on the following eligibility criteria:

### Study designs

Only randomized controlled trials will be included. Non-randomized clinical trials, observational studies (cohort, case-control, and cross-sectional studies), case series, and case reports will be excluded.

### Participants

Parents or guardians of healthy children aged 0 to 12 years who are the direct recipients of the interventions aimed at improving children ‘eating behaviours will be included. Studies targeting children with overweight or obesity will be eligible, however, studies focusing exclusively on children with medical conditions, such as diabetes and neurodevelopmental disorders, will be excluded. Interventions with indirect parental involvement (e.g., provision of information such as newsletters without active engagement) will be excluded.

### Interventions

Any intervention that directly targets children’s eating behaviours and actively involves parents or caregivers will be eligible. Interventions can be single- or multi-component (i.e., employing more than one approach to impact child eating behaviour), provided that eating behaviours are measured as a primary or secondary outcome.

### Comparators

Eligible comparators include no intervention, delayed intervention (wait-list control), usual care, or an alternative intervention that does not influence eating behaviours as the control (e.g. physical activity-only programs).

### Outcomes

The primary outcome will be changes in children’s eating behaviours, encompassing both food-approach and food-avoidant behaviours, as assessed through validated tools or direct observation. Secondary outcomes will include dietary intake, anthropometric measures, quality of life, and parental feeding practices.

### Timing

There will be no restrictions on the duration of the intervention or length of follow-up.

### Setting

Interventions will be included regardless of delivery setting (e.g., home, school, healthcare, or community), provided that parents or caregivers were the primary recipient of the intervention or were actively engaged in its delivery.

### Language

We will include articles reported in English, along with non-English studies that can be adequately translated using Google Translate.

### Information sources

We will search electronic databases from inception to September 2025: MEDLINE (Ovid), EMBASE (Ovid), the Cochrane Central Register of Controlled Trials (CENTRAL) (Ovid), APA PsycINFO (Ovid), Cumulative Index to Nursing and Allied Health Literature (CINAHL) (EBSCO), Scopus, and Web of Science. No date or language restrictions will be applied. The search will be updated at the end of the review to identify newly published or in-press studies prior to the final analysis. To complement our systematic literature search, we will search grey literature (e.g., ClinicalTrials.gov, WHO International Clinical Trials Registry Platform), OpenAire, and other relevant grey literature sources. We will also scan the reference lists of included studies or relevant reviews identified through the search process. Where applicable, we will search the articles’ supplementary materials to make sure that all relevant data will be captured.

### Search strategy

In collaboration with a health information specialist (EU), a comprehensive MEDLINE search strategy was developed using a combination of subject headings and text words related to parent AND children AND eating behaviours and then adapted to other databases. The complete search strategies for each database are provided in **supplementary data 2**.

### Study records

#### Selection process

Two authors will independently screen titles and abstracts of all identified records using the Covidence software (16). Prior to the formal screening process, a pilot exercise will be conducted to ensure the consistency and precision of the screening questions. Subsequently, the full-text articles of the selected records will undergo review by the same authors to determine their eligibility for inclusion. If there are any discrepancies between reviewers at any stage of the screening process that cannot be resolved through consensus, a third reviewer will be consulted to facilitate decision making. Cohen’s kappa will be calculated to assess interrater agreement. We will contact authors if study information required for inclusion determination is unavailable or ambiguous. Reasons for excluding any full-text manuscripts will be documented at this stage, and we will record the selection process in adequate detail to complete a PRISMA flow diagram (17).

#### Data collection process

Pairs of independent reviewers will extract data using a piloted, standardized data extraction form developed by the review team. For included studies, the following will be extracted: a) Study characteristics—first author, publication year, country, study design, setting, sample size; b) Participant characteristics (parent and child)—age, sex, weight status; c) Intervention characteristics —name of the program, intervention description, duration (in weeks), and intensity (number of hours or sessions); (d) comparator characteristics—setting, timing, duration and intensity; and e) Outcomes —study results relevant to our primary outcomes (eating behaviour domains), the tools used to measure them and secondary outcomes.

Any discrepancies between reviewers will be mediated by a third reviewer. If critical information needed for data extraction is unavailable, we will reach out to the corresponding authors a maximum of two times (i.e., initial and follow-up) to obtain additional details.

#### Outcomes and prioritization

The primary outcome will include various aspects of eating behaviours, whether measured by psychometric questionnaires or direct observations in laboratory settings. Measures of child eating behaviour may include but are not limited to: (a) food approach behaviours (e.g., eating in the absence of hunger, enjoyment of food, and desire to drink); and (b) food avoidance behaviours (e.g., satiety responsiveness, slowness in eating and food neophobia). Definitions for the eating behaviour and its domains are provided in **supplementary data 3**.

The secondary outcomes were selected based on current literature identifying factors that influence or are influenced by children’s eating behaviours (1,18). These outcomes include:

1. **Dietary intake** measured using validated methods (e.g., 24-hour recalls, food frequency questionnaires), including macro- and micronutrient intake, food group consumption, and specific dietary components (e.g., sugar, sugar-sweetened beverages).
2. **Anthropometric measures** (parent-reported or measured by trained staff), including absolute body weight, Body Mass Index (BMI/zBMI score), waist circumference, and body composition measures such as percent body fat (via skinfold thickness, bioelectrical impedance analysis, or dual-energy x-ray absorptiometry).
3. **Quality of life** assessed with validated, generic tools (e.g., Pediatric Quality of Life Inventory(19)).
4. **Parental Feeding practices** assessed using validated questionnaires (e.g., Child Feeding Questionnaire(20)).

Secondary outcomes will only be extracted when at least one eligible primary outcome is reported, to ensure the review remains focused on eating behaviours as the primary endpoint.

#### Assessment of risk of bias

Two reviewers will be independently assessing the risk of bias of individual studies, using the Revised Cochrane risk-of-bias tool for randomized trials (RoB2)(21). The RoB2 (individual RCT) consists of five domains focusing on randomisation, deviations from intended interventions, missing outcome data, outcome measurement and in selection of the reported result. One more domain, namely ‘Timing of identification or recruitment of participants,’ was included in the RoB2 (cluster RCT). The questions included in each domain will be answered according to five response options (‘yes,’ ‘probably yes,’ ‘probably no,’ ‘no,’ and ‘no information’). Each domain is judged as either ‘low risk,’ ‘some concerns,’ or ‘high risk,’ based on the answers to the questions, which further results in a proposed overall risk judgment for the specific study being examined. Where required, a third review author will adjudicate discrepancies regarding RoB2 that could not be resolved via consensus.

#### Data synthesis and analysis

A narrative synthesis of study and intervention characteristics will be conducted for all included studies. Continuous data will be reported as means with standard deviations and categorical data as counts and percentages. When examining the characteristics associated with effective interventions, programs are deemed effective if the authors reported a significant difference (at p < 0.05) between the intervention and control groups on at least one outcome. Pooled estimates of effect sizes will be presented as risk ratios with 95% confidence intervals (CIs) for dichotomous variables and mean difference (MD) with 95% CIs for continuous variables. If different measurement scales are used between studies, a standardized MD with 95% CIs will be presented. Provided there are at least two studies, we plan to pool measures of the same quantitative outcomes using the appropriate random effect model. Forest plots will be used to display results.

For studies reporting eating behaviour outcomes at multiple time points (e.g., 6-month and 12-month follow-up), we will group outcomes into predefined follow-up periods (e.g., short-term: ≤6 months; long-term: >6 months) to ensure comparability across studies. If studies report outcomes at unequal intervals, we will assign the closest time point to the predefined categories. For studies with multiple time points, the primary analysis will focus on the time point closest to the predefined primary follow-up period (e.g., 6 months) to maintain consistency, with secondary analyses exploring other time points. Unit of analysis issues will be addressed according to the guidance outlined in the Cochrane Handbook for Systematic Reviews of Interventions(22). For cluster RCTs, we will adjust for clustering using robust standard errors (SEs). For cross-over trials, only data from the first phase will be included. For multi-arm trials, we will combine or appropriately split arms in line with Cochrane Handbook guidance to avoid double-counting, and arms irrelevant to the review question will be dropped. Where necessary, standard deviations will be imputed from SEs, confidence intervals, or p-values. Statistical analysis will be performed using R software version 4.4.0 or greater(23). Comparisons will be 2-tailed using a threshold of p < 0.05 for significance.

#### Missing data

If there is important data missing that is required for analysis, we will attempt to contact the corresponding authors of the studies. If we are unable to resolve the issue, we will proceed with an analysis based on the available data and discuss the potential impact of the missing information. Evidence of potential reporting bias will be documented in the ‘Risk of Bias’ table.

#### Assessment of heterogeneity

Heterogeneity will be evaluated using forest plots and visual assessment of funnel plots for asymmetry. In addition, we will quantify statistical heterogeneity by conducting an X^2^ test and calculating the I^2^ statistic (0% to 40%: might not be important; 30% to 60%: may represent moderate heterogeneity; 50% to 90%: may represent substantial heterogeneity; 75% to 100%: considerable heterogeneity)(24). Study heterogeneity will be informed by a narrative description of study characteristics and causes for study heterogeneity will be explored by subgroup and sensitivity analyses.

#### Subgroup analysis

If sufficient data is available, we plan to conduct analysis across subgroups related to population age (early childhood [<6 years] vs. middle childhood [6-12 years]), intervention (parents only vs. parent-child dyads), method of delivery (face-to-face vs. virtual), income level of study population (low vs. high), and study follow-up (short-term (<6 months) vs. long-term follow-up (>6 months)).

#### Sensitivity analysis

Sensitivity analysis will be performed to explore the source of heterogeneity by excluding data from studies classified as a high risk of bias.

#### Meta-biases

Information from trial registers and protocols will be compared with published reports to determine the extent of reporting bias. We will also evaluate publication bias through visual inspection of funnel plots and Egger’s regression asymmetry test to assess small study effects, provided there are at least 10 studies included(25).

#### Confidence in cumulative evidence

Grading of Recommendations, Assessment, Development and Evaluation (GRADE) will be used to assess the overall certainty of the available evidence for outcome as recommended by the Cochrane handbook(26). Based on our GRADE assessment, the quality of evidence will be assessed across the domains of risk of bias, consistency, directness, precision, and publication bias. Our level of certainty will be presented as either high (further research is very unlikely to change our confidence in the estimate of effect), moderate (further research is likely to have an important impact on our confidence in the estimate of effect and may change the estimate), low (further research is very likely to have an important impact on our confidence in the estimate of effect and is likely to change the estimate), or very low (very uncertain about the estimate of effect).

#### Ethics and dissemination

Ethical approval is not required for this study as individual patient data will not be collected or analyzed. The results will be submitted for publication in a peer-reviewed journal and reported in accordance with the PRISMA 2020 guidelines. Findings will also be shared with researchers, clinicians, and public health stakeholders through academic conferences and relevant knowledge translation activities.

#### Patient and public involvement

Patient partners were involved in identifying the research topic as a priority. They will also be engaged in the dissemination of findings to relevant audiences.

## Discussion

Poor eating behaviours developed in early childhood can lead to both immediate and long-term health consequences. Parents play a crucial role in shaping children’s eating behaviours by promoting practices and strategies that reinforce the behaviours they consider appropriate for their children. It is essential to identify practical guidance and strategies that help provide parents with the skills needed to establish healthy eating behaviours in their children during these crucial years and promote continued positive eating behaviours into adulthood. This systematic review aims to provide an up-to-date synthesis of interventions actively involved parents and assess their impact on child eating behaviours, anthropometrics, diet as well as other related health outcomes, including quality of life and parental feeding practices.

Given the growing interest in implementing interventions targeted at parents to improve children’s health behaviours in early childhood, the findings of this study will provide valuable insights into the effectiveness of these approaches. By identifying the key characteristics of effective interventions, this research will be highly relevant to researchers and community health providers who design and deliver behaviour change programs aimed at families and fostering healthier eating behaviours in children.

## Supporting information

Supplementary material 1-PRISMA-P-SystRev-checklist

Supplementary material 2-Search Strategy

Supplementary material 3-Definition of eating behaviour and related terms

## Data Availability

All data produced in the present study are available upon reasonable request to the authors.

## Authors’ contributions

FK and CB conceptualized the review and drafted the manuscript. FK and EU developed the search strategy. PP and KT contributed to the data synthesis methodology and advised on the statistical analysis. All authors contributed to defining the eligibility and exclusion criteria, provided revisions to the manuscript, and approved the final version of the protocol. CB is the guarantor of the review.

