## Supplementary material 1-PRISMA-P-SystRev-checklist for "Effectiveness of interventions involving parents on children’s eating behaviours: protocol for a systematic review and meta-analysis"

**PRISMA-P 2015 Checklist**

### **This checklist has been adapted for use with protocol submissions to *Systematic Reviews* from Table 3 in Moher D et al**:**** Preferred reporting items for systematic review and meta-analysis protocols (PRISMA-P) 2015 statement. *Systematic Reviews* 2015 ****4****:1

| **Section/topic** | **#** | **Checklist item** | **Information reported** | | **Line number(s)** |
| --- | --- | --- | --- | --- | --- |
|  |  |  | **Yes** | **No** |  |
| **ADMINISTRATIVE INFORMATION** | | | | | |
| **Title** | | | | | |
| Identification | 1a | Identify the report as a protocol of a systematic review | ☒ | ☐ | 2 |
| Update | 1b | If the protocol is for an update of a previous systematic review, identify as such | ☐ | ☒ |  |
| **Registration** | 2 | If registered, provide the name of the registry (e.g., PROSPERO) and registration number in the Abstract | ☒ | ☐ | 44 |
| **Authors** | | | | | |
| Contact | 3a | Provide name, institutional affiliation, and e-mail address of all protocol authors; provide physical mailing address of corresponding author | ☒ | ☐ | 6-21 |
| Contributions | 3b | Describe contributions of protocol authors and identify the guarantor of the review | ☒ | ☐ | 311-316 |
| **Amendments** | 4 | If the protocol represents an amendment of a previously completed or published protocol, identify as such and list changes; otherwise, state plan for documenting important protocol amendments | ☒ | ☐ | 106-108 |
| **Support** | | | | | |
| Sources | 5a | Indicate sources of financial or other support for the review | ☒ | ☐ | 318-319 |
| Sponsor | 5b | Provide name for the review funder and/or sponsor | ☐ | ☒ |  |
| Role of sponsor/funder | 5c | Describe roles of funder(s), sponsor(s), and/or institution(s), if any, in developing the protocol | ☐ | ☒ |  |
| **INTRODUCTION** | | | | | |
| **Rationale** | 6 | Describe the rationale for the review in the context of what is already known | ☒ | ☐ | 76-88 |
| **Objectives** | 7 | Provide an explicit statement of the question(s) the review will address with reference to participants, interventions, comparators, and outcomes (PICO) | ☒ | ☐ | 96-101 |
| **METHODS** | | | | | |
| **Eligibility criteria** | 8 | Specify the study characteristics (e.g., PICO, study design, setting, time frame) and report characteristics (e.g., years considered, language, publication status) to be used as criteria for eligibility for the review | ☒ | ☐ | 109-145 |
| **Information sources** | 9 | Describe all intended information sources (e.g., electronic databases, contact with study authors, trial registers, or other grey literature sources) with planned dates of coverage | ☒ | ☐ | 146-156 |
| **Search strategy** | 10 | Present draft of search strategy to be used for at least one electronic database, including planned limits, such that it could be repeated | ☒ | ☐ | 157-161 |
| ***STUDY RECORDS*** | | | | | |
| Data management | 11a | Describe the mechanism(s) that will be used to manage records and data throughout the review | ☒ | ☐ | 165 |
| Selection process | 11b | State the process that will be used for selecting studies (e.g., two independent reviewers) through each phase of the review (i.e., screening, eligibility, and inclusion in meta-analysis) | ☒ | ☐ | 163-173 |
| Data collection process | 11c | Describe planned method of extracting data from reports (e.g., piloting forms, done independently, in duplicate), any processes for obtaining and confirming data from investigators | ☒ | ☐ | 174-176, 183-185 |
| **Data items** | 12 | List and define all variables for which data will be sought (e.g., PICO items, funding sources), any pre-planned data assumptions and simplifications | ☒ | ☐ | 176-182 |
| **Outcomes and prioritization** | 13 | List and define all outcomes for which data will be sought, including prioritization of main and additional outcomes, with rationale | ☒ | ☐ | 187-208 |
| **Risk of bias in individual studies** | 14 | Describe anticipated methods for assessing risk of bias of individual studies, including whether this will be done at the outcome or study level, or both; state how this information will be used in data synthesis | ☒ | ☐ | 210-221 |
| ***DATA*** | | | | | |
| **Synthesis** | 15a | Describe criteria under which study data will be quantitatively synthesized | ☒ | ☐ | 231-232 |
|  | 15b | If data are appropriate for quantitative synthesis, describe planned summary measures, methods of handling data, and methods of combining data from studies, including any planned exploration of consistency (e.g., *I* ^2^, Kendall’s tau) | ☒ | ☐ | 227-247, 254-261 |
|  | 15c | Describe any proposed additional analyses (e.g., sensitivity or subgroup analyses, meta-regression) | ☒ | ☐ | 262-269 |
|  | 15d | If quantitative synthesis is not appropriate, describe the type of summary planned | ☒ | ☐ | 223-226 |
| **Meta-bias(es)** | 16 | Specify any planned assessment of meta-bias(es) (e.g., publication bias across studies, selective reporting within studies) | ☒ | ☐ | 270-274 |
| **Confidence in cumulative evidence** | 17 | Describe how the strength of the body of evidence will be assessed (e.g., GRADE) | ☒ | ☐ | 275-284 |
