## Supplementary material 2-Search Strategy for "Effectiveness of interventions involving parents on children’s eating behaviours: protocol for a systematic review and meta-analysis"

### Supplementary data 3.

### Search strategy in OVID MEDLINE

| # | Searches |
|---|---|
| 1 | feeding behavior/ |
| 2 | ((feeding or diet or dietary or food) adj3 (behaviour* or behavior* or habit or habits)).ti,ab,kf. |
| 3 | (eating or overeating or undereating or meal or meals or mealtime or satiety or satiation or appetitive trait* or (desire adj1 drink*)).ti,ab,kf. |
| 4 | food fussiness/ |
| 5 | ((food or eater*) adj3 (picky or fussy or fussiness*)).ti,ab,kf. |
| 6 | satiation/ or satiety response/ |
| 7 | appetite/ |
| 8 | or/1-7 [*****Food preference terms*****] |
| 9 | Parents/ or Parenting/ or Legal Guardians/ or Mothers/ or Fathers/ or Grandparents/ or Family/ |
| 10 | (parent*3 or guardian* or mum or mom or mother* or father* or dad or grandparent* or grandmother* or grandfather* or famil*).ti,ab,kf. |
| 11 | exp parent-child relations/ |
| 12 | or/9-11 [******parent term*****] |
| 13 | (intervention* or intervene* or input or influence* or behavior* or behaviour* or preference* or selection* or selecting or select or selects or buy or buys or choice* or purchas*).ti,ab,kf. |
| 14 | 12 and 13 |
| 15 | 8 and 14 |
| 16 | limit 15 to ("all infant (birth to 23 months)" or "preschool child (2 to 5 years)" or "child (6 to 12 years)") |
| 17 | (infan* or child* or toddler* or pre-school* or preschool* or school-age* or youth* or pre-adolescent* or paediatric* or pediatric* or boy or boys or girl or girls or minor* or juvenile* or pubescen* or prepubescen* or kindergarten).ti,ab,kf. |
| 18 | 16 or (15 and 17) [*****age group limit****] |
| 19 | randomized controlled trial.pt. |
| 20 | controlled clinical trial.pt. |
| 21 | ((randomised or randomized or control*) adj2 trial).ti,ab,kf. |
| 22 | randomized.ab. |
| 23 | placebo.ab. |
| 24 | randomly.ab. |
| 25 | trial.ab. |

| | |
|---|---|
| 26 | groups.ab. |
| 27 | or/19-26 [****Clincial trial terms****] |
| 28 | 18 and 27 [**Final Results**] |

### Search strategy in OVID EMBASE

| # | **Searches** |
|---|---|
| 1 | feeding behavior/ |
| 2 | ((feeding or diet or dietary or food) adj3 (behavior* or behaviour* or habit or habits)).ti,ab,kf. |
| 3 | (eating or overeating or undereating or meal or meals or mealtime or satiety or satiation or appetitive trait* or (desire adj1 drink*)).ti,ab,kf. |
| 4 | food fussiness/ |
| 5 | ((food or eater*) adj3 (picky or fussy or fussiness*)).ti,ab,kf. |
| 6 | satiation/ or satiety response/ |
| 7 | (satiation or satiety).ti,ab,kf. |
| 8 | appetite/ or eating habit/ or meal size/ or portion size/ or sodium appetite/ |
| 9 | (appetite or ((portion or meal) adj2 size*)).ti,ab,kf. |
| 10 | or/1-9 [*****Food preference terms*****] |
| 11 | parent/ or father/ or adolescent father/ or mother/ or adolescent mother/ or single parent/ or grandparent/ or grandfather/ or grandmother/ |
| 12 | (parent*3 or guardian* or mum or mom or mother* or father* or dad or grandparent* or grandmother* or grandfather* or famil*).ti,ab,kf. |
| 13 | family/ or parenthood/ or single-parent family/ or legal guardian/ |
| 14 | child parent relation/ or father child relation/ or mother child relation/ |
| 15 | or/11-14 [******parent term*****] |
| 16 | (intervention* or intervene* or input or influence* or behavior* or behaviour* or preference* or selection* or selecting or select or selects or buy or buys or choice* or purchas*).ti,ab,kf. |
| 17 | 10 and 15 and 16 [****Base set 1 - food preferences and parents and interventions****] |
| 18 | limit 17 to (infant <to one year> or child <unspecified age> or preschool child <1 to 6 years> or school child <7 to 12 years>) |
| 19 | (infan* or child* or toddler* or pre-school* or preschool* or school-age* or youth* or pre-adolescent* or paediatric* or pediatric* or boy or boys or girl or girls or minor* or juvenile* or pubescen* or prepubescen* or kindergarten).ti,ab,kf. |
| 20 | 18 or (17 and 19) [*****age group limit****] |
| 21 | randomized controlled trial/ |

| | |
|---|---|
| 22 | Controlled clinical study/ |
| 23 | random*.ti,ab. |
| 24 | Randomization/ |
| 25 | Intermethod comparison/ |
| 26 | placebo*.ti,ab. |
| 27 | (compare or compared or comparison).ti. |
| 28 | ((evaluated or evaluate or evaluating or assessed or assess) and (compare or compared or comparing or comparison)).ab. |
| 29 | (open adj label).ti,ab. |
| 30 | ((double or single or doubly or singly) adj (blind or blinded or blindly)).ti,ab. |
| 31 | double-blind procedure/ |
| 32 | Parallel group$1.ti,ab. |
| 33 | (Crossover or cross over).ti,ab. |
| 34 | ((assign* or match or matched or allocation) adj5 (alternate or group$1 or intervention$1 or patient$1 or subject$1 or participant$1)).ti,ab. |
| 35 | (assigned or allocated).ti,ab. |
| 36 | (controlled adj7 (study or design or trial)).ti,ab. |
| 37 | (volunteer or volunteers).ti,ab. |
| 38 | Human experiment/ |
| 39 | Trial.ti. |
| 40 | or/21-39 |
| 41 | (Random$ adj sample$ adj7 ("cross section$" or questionnaire$1 or survey$1 or database$1)).ti,ab. not (comparative study/ or controlled study/ or randomi$ed controlled.ti,ab. or randomly assigned.ti,ab.) |
| 42 | Cross-sectional study/ not (randomized controlled study/ or controlled clinical study/ or controlled study/ or randomi?ed controlled.ti,ab. or control groups$1.ti,ab.) |
| 43 | (((case adj control$) and random$) not randomi?ed controlled).ti,ab. |
| 44 | (Systematic review not (trial or study)).ti. |
| 45 | (nonrandom$ not random$).ti,ab. |
| 46 | "random field$".ti,ab. |
| 47 | (random cluster adj3 sampl$).ti,ab. |
| 48 | (review.ab. and review.pt.) not trial.ti. |
| 49 | ("we searched".ab. and review.ti.) or review.pt. |
| 50 | "update review".ab. |
| 51 | (databases adj4 searched).ab. |

| | |
|---|---|
| 52 | (rat or rats or mouse or mice or swine or porcine or murine or sheep or lambs or pigs or piglets or rabbit or rabbits or cat or cats or dog or dogs or cattle or bovine or monkey or monkeys or trout or marmoset$1).ti. and animal experiment/ |
| 53 | Animal experiment/ not (human experiment/ or human/) |
| 54 | or/41-53 |
| 55 | 40 not 54 [****Cochrane Box 3.e EMBASE sensitive Therapy Treatment Effectiveness Filter terms 2018 revision (Glainville et al 2019b)****] |
| 56 | 20 and 55 [****Final results****] |

### Search strategy in CENTRAL Database

| # | **Searches** |
|---|---|
| 1 | feeding behavior/ |
| 2 | ((feeding or diet or dietary or food) adj3 (behavior* or behaviour* or habit or habits)).ti,ab,kf. |
| 3 | (eating or overeating or undereating or meal or meals or mealtime or satiety or satiation or appetitive trait* or (desire adj1 drink*)).ti,ab,kf. |
| 4 | food fussiness/ |
| 5 | ((food or eater*) adj3 (picky or fussy or fussiness*)).ti,ab,kf. |
| 6 | satiation/ or satiety response/ |
| 7 | (satiation or satiety).ti,ab,kf. |
| 8 | appetite/ |
| 9 | or/1-8 |
| 10 | Parents/ or Parenting/ or Legal Guardians/ or Mothers/ or Fathers/ or Grandparents/ or Family/ |
| 11 | (parent*3 or guardian* or mum or mom or mother* or father* or dad or grandparent* or grandmother* or grandfather* or famil*).ti,ab,kf. |
| 12 | exp parent-child relations/ |
| 13 | or/10-12 |
| 14 | (intervention* or intervene* or input or influence* or behavior* or behaviour* or preference* or selection* or selecting or select or selects or buy or buys or choice* or purchas*).ti,ab,kf. |
| 15 | 13 and 14 |
| 16 | 9 and 15 |
| 17 | (infan* or child* or toddler* or pre-school* or preschool* or school-age* or youth* or pre-adolescent* or paediatric* or pediatric* or boy or boys or girl or girls or minor* or juvenile* or pubescen* or prepubescen* or kindergarten).ti,ab,kf. |
| 18 | 16 and 17 [***Final Results***] |

### Search strategy in CINAHL

| # | **Searches** |
|---|---|
| 1 | (MH "Eating Behavior") OR (MH "Food Fussiness") OR (MH "Appetite") OR (MH "Satiation") |
| 2 | TI ( ((feeding or diet or dietary or food) N3 (behavior* or behaviour* or habit or habits)) ) OR AB ( ((feeding or diet or dietary or food) N3 (behavior* or behaviour* or habit or habits)) ) |
| 3 | TI ( (eating or overeating or undereating or meal or meals or mealtime or satiety or satiation or appetitive trait* or (desire N1 drink*)) ) OR AB ( (eating or overeating or undereating or meal or meals or mealtime or satiety or satiation or appetitive trait* or (desire N1 drink*)) ) |
| 4 | TI ( ((food or eater*) N3 (picky or fussy or fussiness*)) ) OR AB ( ((food or eater*) N3 (picky or fussy or fussiness*)) ) |
| 5 | S1 OR S2 OR S3 OR S4 |
| 6 | (MH "Parents") OR (MH "Fathers") OR (MH "Adolescent Fathers") OR (MH "Mothers") OR (MH "Adolescent Mothers") OR (MH "Mothers, Working") OR (MH "Single Parent") OR (MH "Family") OR (MH "Grandparents") OR (MH "Extended Family") OR (MH "Parenting") OR (MH "Guardianship, Legal") |
| 7 | TI ( (parent* or guardian* or mum or mom or mother* or father* or dad or grandparent* or grandmother* or grandfather* or famil*) ) OR AB ( (parent* or guardian* or mum or mom or mother* or father* or dad or grandparent* or grandmother* or grandfather* or famil*) ) |
| 8 | (MH "Parent-Child Relations") OR (MH "Father-Child Relations") OR (MH "Mother-Child Relations") OR (MH "Parent-Infant Relations") OR (MH "Father-Infant Relations") OR (MH "Mother-Infant Relations") |
| 9 | S6 OR S7 OR S8 |
| 10 | TI ( (intervention* or intervene* or input or influence* or behavior* or behaviour* or preference* or selection* or selecting or select or selects or buy or buys or choice* or purchas*) ) OR AB ( (intervention* or intervene* or input or influence* or behavior* or behaviour* or preference* or selection* or selecting or select or selects or buy or buys or choice* or purchas*) ) |
| 11 | S5 AND S9 AND S10 |
| 12 | S5 AND S9 AND S10 |
| 13 | (MH "Randomized Controlled Trials") OR (MH "Clinical Trials") OR (MH "Intervention Trials") OR (MH "Preventive Trials") |
| 14 | TI (randomized N2 trial) OR AB (randomized N2 trial) |
| 15 | S13 OR S14 |
| 16 | S12 AND S15[***Final Results***] |

### Search strategy in APA PsycInfo

| | Searches |
|---|---|
| 1 | eating behavior/ or dietary restraint/ or emotional eating/ or food refusal/ or healthy eating/ or appetite/ or "avoidant/restrictive food intake disorder"/ or eating attitudes/ or mealtimes/ |
| 2 | ((feeding or diet or dietary or food) adj3 (behavior* or behaviour* or habit or habits)).ti,ab,id. |
| 3 | (eating or overeating or undereating or meal or meals or mealtime or satiety or satiation or appetitive trait* or (desire adj1 drink*)).ti,ab,id. |
| 4 | ((food or eater*) adj3 (picky or fussy or fussiness*)).ti,ab,id. |
| 5 | eating attitudes/ |
| 6 | satiation/ or appetite/ |
| 7 | (satiation or satiety).ti,ab,id. |
| 8 | appetite/ or hunger/ or satiation/ or dietary restraint/ or eating attitudes/ or eating behavior/ |
| 9 | (appetite or ((portion or meal) adj2 size*)).ti,ab,id. |
| 10 | or/1-9 [*****Food preference terms*****] |
| 11 | parents/ or fathers/ or mothers/ or single parents/ or parenting/ |
| 12 | (parent*3 or guardian* or mum or mom or mother* or father* or dad or grandparent* or grandmother* or grandfather* or famil*).ti,ab,id. |
| 13 | family/ or parenthood/ or single-parent family/ or legal guardian/ |
| 14 | child parent relation/ or father child relation/ or mother child relation/ |
| 15 | or/11-14 [******parent term*****] |
| 16 | 10 and 15 |
| 17 | limit 16 to "0300 clinical trial" |
| 18 | randomized clinical trials/ |
| 19 | trial.ab. |
| 20 | random.ab. |
| 21 | or/18-20 |
| 22 | 17 or (16 and 21) |
| 23 | limit 22 to 100 childhood <birth to age 12 yrs> |
| 24 | (infan* or child* or toddler* or pre-school* or preschool* or school-age* or youth* or pre-adolescent* or paediatric* or pediatric* or boy or boys or girl or girls or minor* or juvenile* or pubescen* or prepubescen* or kindergarten).ti,ab,id. |
| 25 | 23 or (22 and 24)[***Final Results***] |

### Search strategy in Web of science

(((((((TS=(((feeding or diet or dietary or food) NEAR/3 (behavior* or behaviour* or habit or habits)))) OR TS=(((food or eater*) NEAR/3 (picky or fussy or fussiness*)))) OR TS=((eating or overeating or undereating or meal or meals or mealtime or satiety or satiation or appetitive trait* or (desire NEAR/1 drink*)).)) AND TS=((parent* or guardian* or mum or mom or mother* or father* or dad or grandparent* or grandmother* or grandfather* or famil*).))) AND TS=((intervention* or intervene* or input or influence* or behavior* or behaviour* or preference* or selection* or selecting or select or selects or buy or buys or choice* or purchas*).)) AND TS=((randomized NEAR/2 trial))) AND TS=((infan* or child* or toddler* or pre-school* or preschool* or school-age* or youth* or pre-adolescent* or paediatric* or pediatric* or boy or boys or girl or girls or minor* or juvenile* or pubescen* or prepubescen* or kindergarten).)

### Search strategy in Scopus

( ( TITLE-ABS-KEY ( feeding OR diet OR dietary OR food W/3 ( behavior* OR behaviour* OR habit OR habits ) ) ) OR ( TITLE-ABS-KEY ( food OR eater* W/3 ( picky OR fussy OR fussiness* ) ) ) OR ( TITLE-ABS-KEY ( eating OR overeating OR undereating OR meal OR meals OR mealtime OR satiety OR satiation OR appetitive AND trait* ) ) OR ( TITLE-ABS-KEY ( desire W/2 drink* ) ) ) AND ( TITLE-ABS-KEY ( parent* OR guardian* OR mum OR mom OR mother* OR father* OR dad OR grandparent* OR grandmother* OR grandfather* OR famil* ) ) AND ( TITLE-ABS-KEY ( intervention* OR intervene* OR input OR influence* OR behavior* OR behaviour* OR preference* OR selection* OR selecting OR select OR selects OR buy OR buys OR choice* OR purchas* ) ) AND ( TITLE-ABS-KEY ( infan* OR child* OR toddler* OR pre-school* OR preschool* OR school-age* OR youth* OR pre-adolescent* OR paediatric* OR pediatric* OR boy OR boys OR girl OR girls OR minor* OR juvenile* OR pubescen* OR prepubescen* OR kindergarten ) ) AND ( TITLE-ABS-KEY ( randomized W/2 trial ) )
