## Supplementary material 3-Definition of eating behaviour and related terms for "Effectiveness of interventions involving parents on children’s eating behaviours: protocol for a systematic review and meta-analysis"

**Supplementary data 2.**

**Definitions and assessment methods of eating behaviour and related constructs in Children**

| Eating behaviour terms | Definition | Method of assessment |
|---|---|---|
| Eating behaviours(1) | A set of predispositions and tendencies toward foods that reflect aspects such as hunger, appetite, satiety, and response to food cues. | |
| ***Food approach behaviours*** | Behaviours and thoughts that involve a movement toward or desire for food | |
| Food responsiveness(2)/ External eating(3) | Eating in response to external (usually food related stimuli) | CEBQ, DEBQ |
| Enjoyment of food(2) | The extent to which palatable foods provoke eating | CEBQ, |
| Emotional overeating(2)/ Emotional eating(3)/ Disinhibition(4) | Overeating in response to negative emotional states | CEBQ, DEBQ, TFEQ |
| Eating in the absence of hunger (EAH)(5) | Eating when satiated in response to the presence of palatable snack food | Direct observation in a laboratory setting |
| Desire to drink(2) | The tendency to carry drinks (often sweetened beverages) | CEBQ |
| Eating rate(6) | Total energy or mouthfuls of food consumed within a given time interval | Direct observation of (usually) videotapes |
| Compensation of energy intake(7) | Adjustments in intake in response to changes in the caloric content of a preload (fixed amount of food or nutrient) after a predetermined time delay | Compensation trials |
| ***Food avoidance behaviours*** | Behaviours and thoughts that involve a movement away from food | |
| Satiety responsiveness(2) | The extent to which children avoid eating, and for how long after, satiation | CEBQ, |
| Food neophobia(8) | Fear or reluctance to consume or an unwillingness to try new or unknown foods. | CFNS |
| Picky/fussy eating(2) | Selectivity regarding which foods are consumed | CEBQ |
| Slowness in eating(2) | Fewer bites per minute, usually as a meal progresses | CEBQ, |

| | | |
|---|---|---|
| Emotional undereating(2) | The extent of a tendency reduces food intake in response to negative emotions | CEBQ |
| Restrained eating(3)/ cognitive restraint of eating(4) | How strong attempts to restrain eating are | DEBQ, TFEQ |
| Hunger(4) | Responsiveness to internal and external hunger sensations | TFEQ |

CEBQ, Child Eating Behaviour Questionnaire; DEBQ, Dutch Eating Behaviour Questionnaire; TFEQ, Three Factor Eating Behaviour Questionnaire; CFNS, Children's Food Neophobia Scale.
